# Measles, Rubella, and Mumps in Mexico: A National Serosurvey Highlighting Reemergence Risks

**DOI:** 10.64898/2026.02.19.26346647

**Authors:** Salas-Lais Angel Gustavo, Fernandes-Matano Larissa, Torres-Flores Alejandro, Morales-Hernández María Luisa, López-Macías Constantino, Martínez-Miguel Bernardo, Tepale-Segura Araceli, Guerrero-García José de Jesús, Alvarado-Yaah Julio Elias, Anguiano-Hernández Yu-Mei, Castro-Escamilla Octavio, Zamudio-Chávez Óscar, Herrera-Gómez Felipe de Jesús, Krug-Llamas Ernesto, Romero-Feregrino Rodrigo, Santacruz-Tinoco Clara Esperanza, C. Bonifaz Laura, Díaz-Jiménez Carlos, Vargas-García Alejandro Manuel, Muñoz-Medina José Esteban, Santos-Carrillo Alva Alejandra

**Author notes:** **Corresponding author** José Esteban Muñoz Médina, Ph.D., Biobank Manager; Division of Specialized Laboratories, Mexican Social Security Institute, 621 Riobamba St., Mexico City 07760. These authors contributed equally to this work, and their order was randomly assigned (Salas-Lais Angel Gustavo, Fernandes-Matano Larissa).

## Abstract

**Objectives:** Despite the availability of effective vaccines, achieving the seroprevalence thresholds recommended by the World Health Organization (WHO) for eliminating measles, rubella, and mumps remains a public health challenge.

**Methods:** A retrospective, cross-sectional serological survey was conducted, including 9,587 serum samples collected from 31 of the 32 federal entities of Mexico between September and December 2024. IgG antibody levels against measles, rubella, and mumps were quantified using chemiluminescent immunoassays. Seroprevalence was analyzed by age, sex, and geographic region.

**Results:** The overall seroprevalence was 78.3% for measles, 88.6% for rubella, and 81.5% for mumps (p<0.05). Age-stratified analysis revealed significant gaps in immunity against measles and mumps, particularly in the 10–39-year-old group, in which seroprevalence dropped below 70%. In contrast, more consistent protection against rubella was observed across all age groups, although younger children showed lower antibody titers. Regional analysis indicated that only older adults reached the protective threshold against measles in all states.

**Conclusions:** This study demonstrates that current levels of seroprevalence in Mexico do not correspond to the vaccination coverage recommended by the WHO and highlights the urgent need to strengthen vaccination strategies, conduct catch-up campaigns, and carry out continuous seroepidemiological surveillance to maintain elimination goals.

## Introduction

Measles, rubella, and mumps are highly contagious viral diseases that have historically affected mainly children and adolescents, although susceptible adults may also be at risk (1-5). These infections are transmitted through respiratory droplets and, prior to the introduction of the measles-mumps-rubella (MMR) vaccine in the 1970s, they caused millions of deaths worldwide annually. Currently, all three diseases are vaccine preventable, and effective immunization programs have significantly reduced their incidence in many countries.

However, the elimination of these viruses requires sufficiently high levels of herd immunity. It is estimated that vaccination coverage must exceed 95% for measles, ranging between 83% and 86% for rubella and between 88% and 92% for mumps (6-9). Studies have shown that these thresholds are not always met, even in countries with robust health care systems. For instance, in Germany, the seroprevalence of mumps was found to be insufficient, and that of rubella barely reached the minimum levels recommended by the WHO (10).

In Mexico, although measles, rubella, and mumps are included in the national immunization schedule, surveillance efforts focus primarily on symptomatic cases that meet operational case definitions. Between 2019 and 2023, more than 11,000 probable cases of measles or rubella were reported, with 217 confirmed measles cases, most of which were imported. During the same period, more than 16,000 confirmed cases of mumps were documented (11-15). Despite ongoing control efforts, the risk of virus reintroduction and transmission persists, driven by population mobility and gaps in vaccination coverage. This underscores the urgent need to strengthen vaccination strategies and epidemiological surveillance to prevent new cases.

The measles (*Morbillivirus hominis*; MVH), rubella (*Rubivirus rubella*; RVR), and mumps (*Orthorubulavirus parotitidis*; OVP) viruses exhibit distinct biological and clinical characteristics but share high transmission potential and the ability to cause severe complications. MVH is among the most contagious viruses known, with a basic reproduction number (R₀) estimated to be between 12 and 18, and is characterized by efficient infection of the respiratory epithelium and rapid systemic dissemination (16,17). Although only one serotype has been identified, multiple genotypes are currently in circulation (18). OVP has marked tropism for glandular and neural tissues, and while it usually causes mild disease, it can lead to complications such as orchitis, meningitis, pancreatitis, and even permanent deafness (3,19). In the case of RVR, infection during pregnancy can result in severe outcomes, including congenital rubella syndrome (CRS), which involves cardiac, neurological, ocular, and auditory abnormalities (20,21).

Despite the availability of effective vaccines and established immunization schedules, recent outbreaks in populations with access to the MRR or MR vaccines have highlighted the need to monitor population-level immunity and reinforce vaccine coverage. Given the global and regional epidemiological context and the high level of population mobility in Mexico, assessing the status of population immunity against these three viruses is essential. Seroprevalence surveys, such as those conducted in this study, can serve as indirect indicators of vaccination coverage and help guide targeted interventions to strengthen the national immunization program.

## Methods

### Study design

A retrospective cross-sectional serological study was conducted that included all age groups and spanned 31 of Mexico’s 32 states. Between September and December 2024, as part of the activities of the Biobank of the Coordination of Quality of Supplies and Specialized Laboratories (CCILE), 31,537 serum samples were collected anonymously from 34 blood banks and 34 clinical laboratories of the Mexican Social Security Institute (IMSS), distributed throughout Mexico. Samples were not accepted if they met any of the following rejection criteria: lipemic, hemolyzed, contaminated, turbid, spilled, or misidentified. Owing to the anonymous nature of the sampling, no patient-traceable data were requested, and only age, sex, and state of origin were obtained. Subsequently, 9,604 samples were randomly selected for analysis. Even though the number of samples was limited by the amount of available resources (samples and tests), an effort was made to eliminate bias by considering a minimum of 300 samples per state distributed across all age groups. During sample processing, 17 additional samples were excluded because of insufficient volume to simultaneously obtain mumps, rubella, and measles IgG antibody results. Therefore, seroprevalence analyses were performed on 9,587 samples with valid results for all three diagnoses (Figure 1).

**Figure 1.**
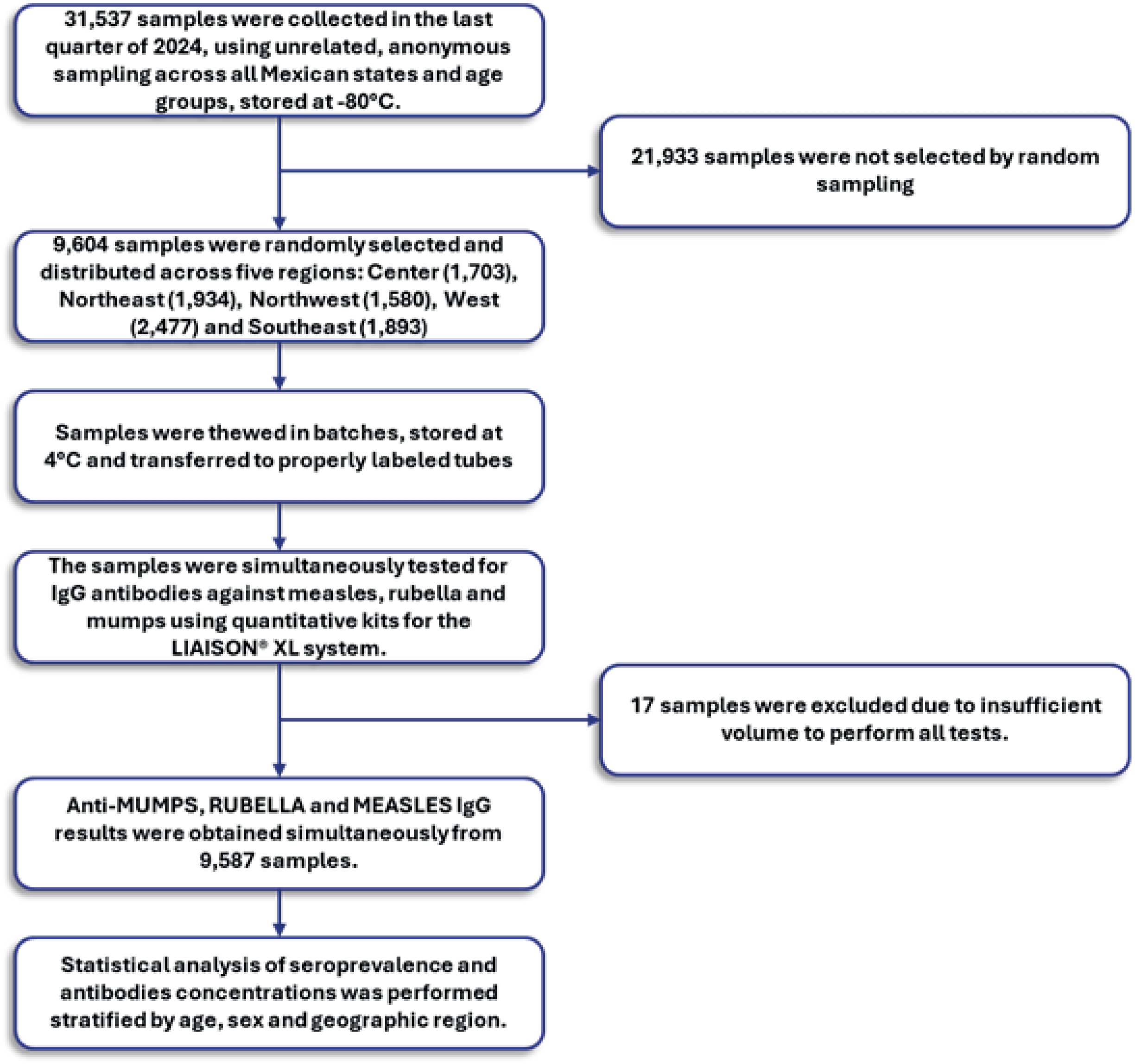
Flowchart of sample selection and analysis. Diagram depicting the study design, the selection process of the analyzed samples, and the laboratory procedures applied.

This study was approved by the Ethics and Research Committees of the National Committee of Scientific Research of the Instituto Mexicano del Seguro Social (registration number R-2025-785-027).

The samples analyzed were from the five regions of Mexico: 1,703 (17.8%) from the Center (Mexico City, State of Mexico, Morelos, Puebla, Tlaxcala, Hidalgo, and Guerrero); 1,934 (20.2%) from the Northeast (Durango, Coahuila, Nuevo León, San Luis Potosí, and Tamaulipas); 1,580 (16.5%) from the Northwest (Baja California, Baja California Sur, Chihuahua, Sonora, and Sinaloa); 2,477 (25.8%) from the West (Aguascalientes, Colima, Guanajuato, Jalisco, Michoacán, Nayarit, Querétaro, and Zacatecas); and 1,893 (19.7%) from the Southeast (Veracruz, Chiapas, Oaxaca, Campeche, Quintana Roo, Tabasco, and Yucatán).

### Measuring IgG antibodies to measles, rubella, and mumps

IgG antibodies against MVH, RVR, and OVP were detected and quantified using a chemiluminescent immunoassay (CLIA) on an automated LIAISON^®^ XL analyzer (DiaSorin, Saluggia, Italy). For each sample, 170 µL of serum (20 µL + 150 µL dead volume) was needed. The following specific commercial kits were used: LIAISON^®^ Measles IgG, LIAISON^®^ Rubella IgG II, and LIAISON^®^ Mumps IgG. The results were interpreted according to the manufacturer’s recommendations: for measles, ≥ 16.5 AU/mL was considered positive, 13.5–16.5 AU/mL was borderline, and < 13.5 AU/mL was negative [specificity (Sp): 97.4%; sensitivity (Se): 94.7%]; for rubella, ≥10 IU/mL was considered positive, 7–10 IU/mL was borderline, and <7 IU/mL was negative (Sp: 99.1%; Se: 99.5%), and for mumps, ≥11.0 AU/mL was considered positive, 9.0–11.0 AU/mL was borderline, and <9.0 AU/mL was negative (Sp: 98.2%; 98.5%). In all cases, calibrators provided by the manufacturer were used to construct standard curves, and internal quality controls were included in each assay.

### Statistical analysis

Seroprevalence values were reported using descriptive statistics, calculated as the proportion of positive cases in relation to the total number of samples analyzed, stratified by age group, sex, and geographic region, and 95% confidence intervals were calculated for each proportion. Comparisons between categorical variables were performed using the chi-square test for homogeneity and independence, as well as the Phi coefficient. For continuous variables, the Kruskal-Wallis test or Mann-Whitney U test was applied because of the nonparametric characteristics of the data. Outliers were identified using the interquartile range method (IQR). A p value < 0.05 was considered to indicate statistical significance in all cases. Analyses and graphing were carried out using IBM SPSS Statistics^®^ (version 27.0; Armonk, NY, USA) and Microsoft^®^ Excel^®^ (version 16.66.1; Redmond, WA, USA) and edited with Adobe Illustrator^®^ (version 25.0; San Jose, CA).

## Results

### Demographic characteristics of the study population

Among the 9,587 samples analyzed, 4,999 (52.1%) were from women, and 4,588 (47.9%) were from men. The dataset included participants from all age groups, namely, children (0-9 years), adolescents (10-19 years), adults (20-59 years), and older adults (≥60 years), providing a solid foundation for comparative analyses by age, sex, and geographic region (Table 1).

**Table 1.**
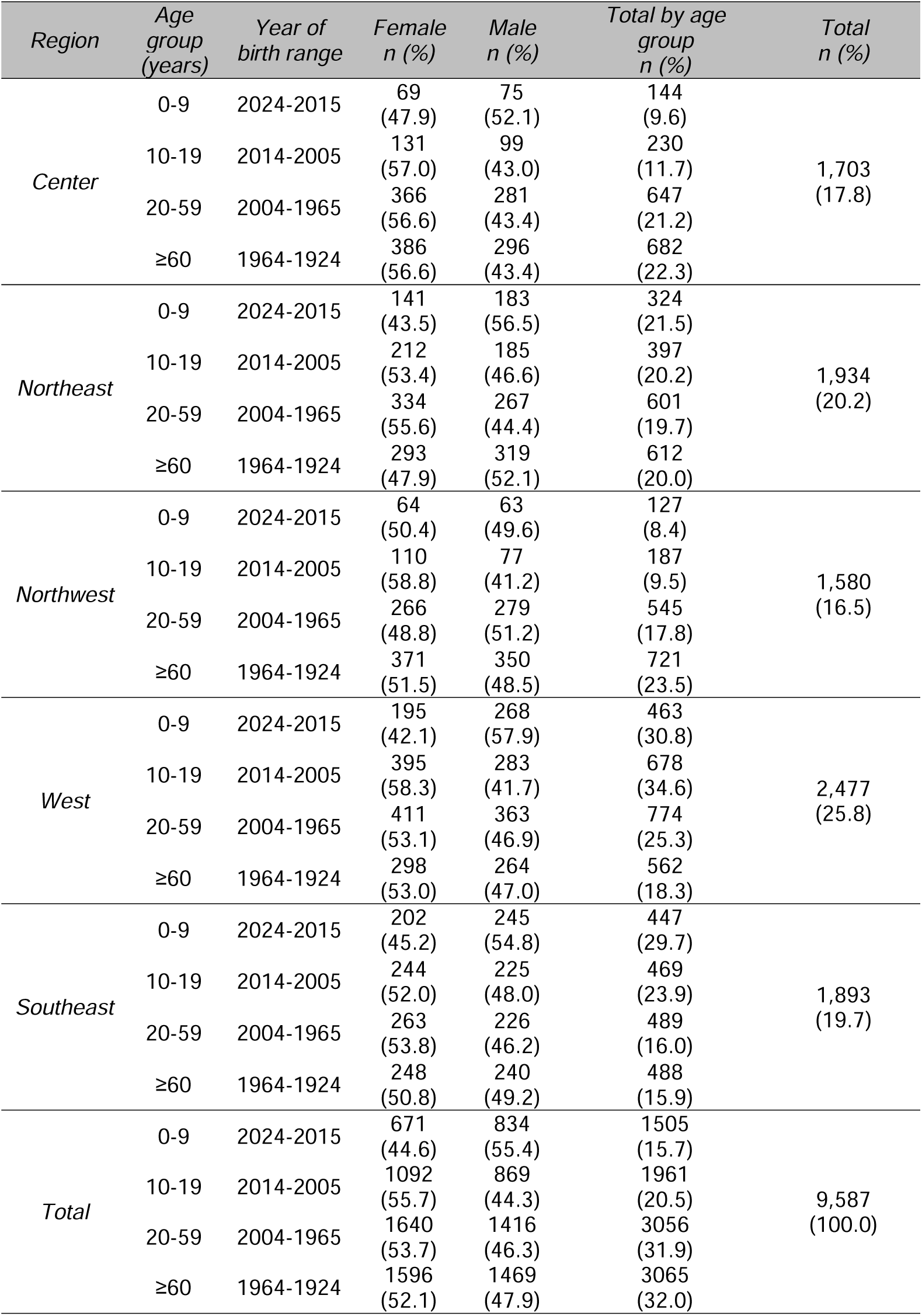
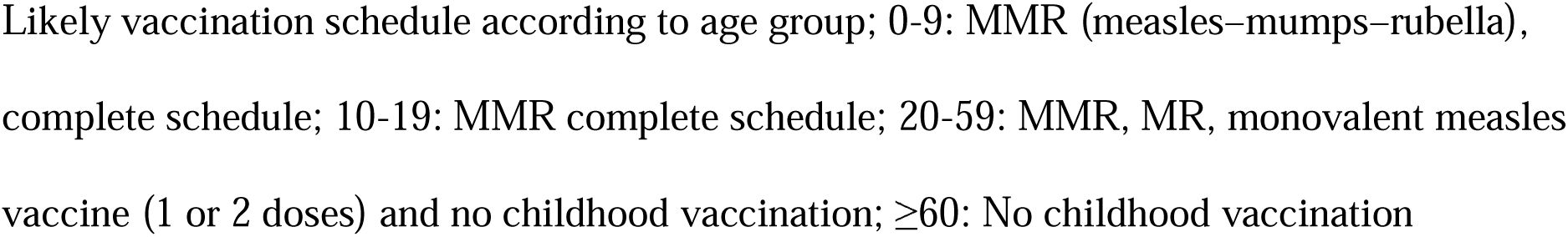
Demographic data by region and age group.

### Seroprevalence of mumps, rubella, and measles

Seroprevalence analysis was performed, stratifying the samples by age group and subgroup (Figure 2). Significant differences were found in the overall seroprevalence for the three different diagnoses: mumps, 81.5%; rubella, 88.6%; and measles, 78.3% (p<0.05). Significant differences were also observed among age groups for the same disease.

**Figure 2.**
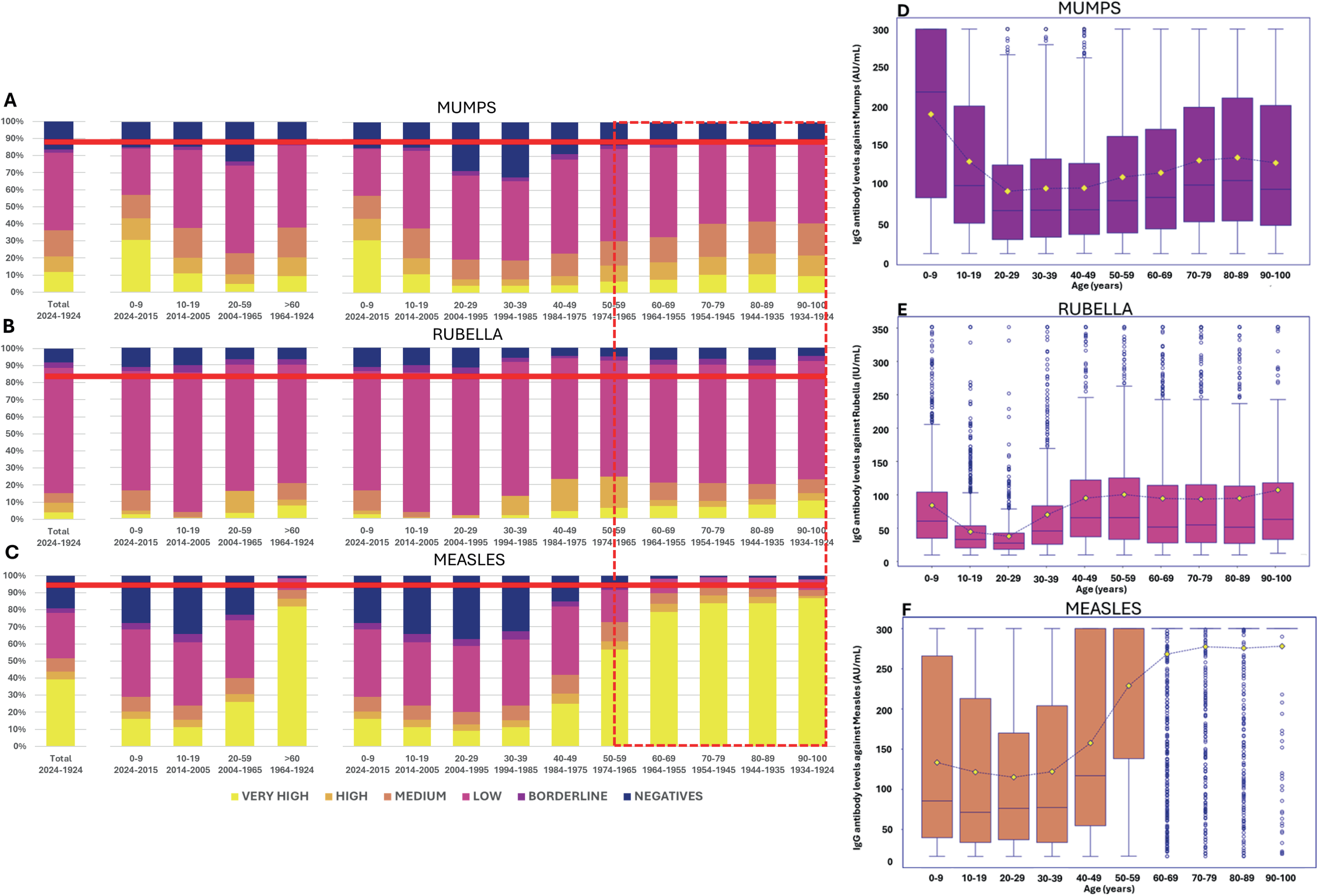
Seroprevalence of mumps, rubella, and measles in Mexico by age group and subgroup. (A–C) Proportion of samples with IgG seropositivity for mumps (A), rubella (B), and measles (C) by age group. The bars show the percentage of positive, borderline, and negative samples (y-axis: percentage of total samples). Red horizontal lines indicate the WHO-recommended seroprevalence thresholds to prevent outbreaks, whereas dashed boxes highlight age groups in which immunity is primarily attributable to natural viral circulation. The overall seroprevalence observed in this study was 81.5% for mumps, 88.6% for rubella, and 78.3% for measles. (D–F) IgG antibody titers for mumps (D), rubella (E), and measles (F) stratified by age subgroup (y-axis: IgG antibody concentration, AU/mL or IU/mL). Yellow dots represent mean values, and horizontal lines within the boxes represent the median.

A lower seroprevalence was observed for mumps among adults aged 20-59 years (mainly in the 20-29 and 30-39 subgroups) than among children and older adults (Figure 2A). Among adults, the seroprevalence was the lowest, and IgG antibody levels were also lower (p<0.05) (Figure 2D). The highest seroprevalence was observed in adults aged ≥ 60 years; however, the highest antibody levels were found in children aged 0-9 years (p<0.05).

As shown in Figure 2C, the seroprevalence of measles varied the most between age groups (p<0.05). The most susceptible subgroups were adolescents aged 10-19 years and adults aged 20-39, with seroprevalence less than 70%. Antibody quantities were also the lowest in these age groups (p<0.05) (Figure 2F). In subgroups aged 40 and over, an increase in both seroprevalence and antibody levels was observed.

In contrast, the rubella seroprevalence was the most consistent between age groups, with the youngest individuals presenting the lowest IgG antibody percentages and concentrations (Figure 2B and E).

In general, the analysis of mumps, rubella, and measles seroprevalence by sex revealed no statistically significant differences. In contrast, analyses of antibody levels revealed statistically significant differences in the mean rubella IgG antibody levels between men and women; however, when the Mann-Whitney U statistic was converted to a z score, the rank biserial correlation (r) was calculated, revealing an almost negligible effect from the small differences observed (mumps r = 0.02, rubella r = 0.07, and measles r = 0.01) (Figure 3).

**Figure 3.**
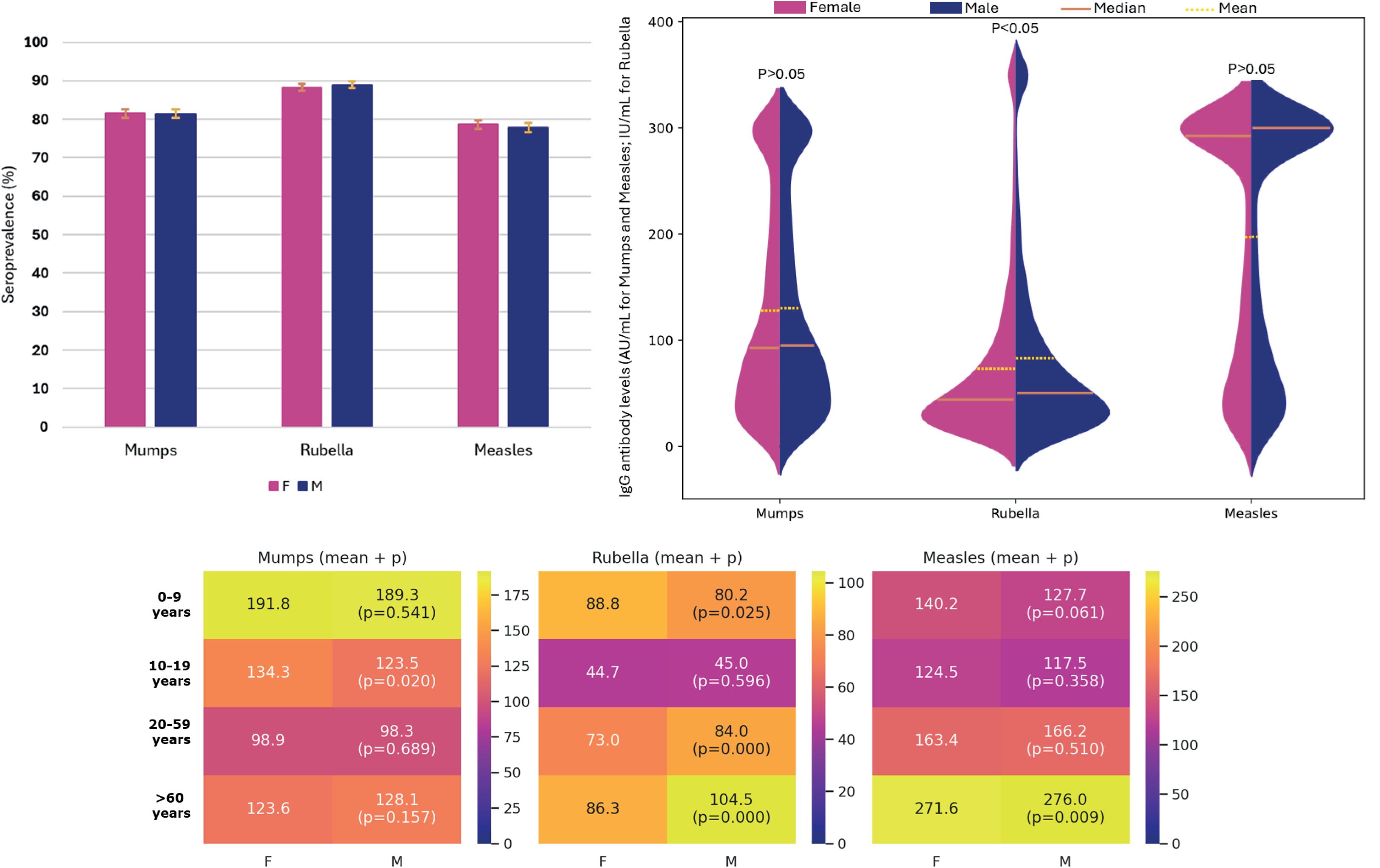
Comparison of seroprevalence and IgG antibody titers against mumps, rubella, and measles by sex. A) Seroprevalence percentages stratified by sex. B) Violin plots showing the distribution of IgG antibody titers against the three viruses by sex; horizontal lines represent the median, and dotted lines indicate the mean. C) Heatmap displaying differences and levels of statistical significance in IgG antibody titers by sex and age group.

To identify risk zones, we performed a seroprevalence analysis by region. The colors shown on the maps in Figure 4 indicate the regions still susceptible to outbreaks, according to the seroprevalence percentages indicated by the WHO to ensure virus elimination for each case (mumps: 88–92%, rubella: 83–86%, and measles: >95%). As shown in Figure 5B, only children aged 0–9 years in the northeast of the country remained susceptible to rubella, with a seroprevalence percentage that differed significantly from that in the other regions (northeast: 77.2% vs. center: 84.7%, northwest: 81.1%, west: 86.4%, and southeast: 87.7%; p<0.05). On the other hand, for mumps (Figure 5A), all maps are fully colored, although the seroprevalence differences between each region are significant only for children and adults. This catches our attention because, among the viruses analyzed, it is the one that presents the most cases per year (although with far fewer repercussions on health), in addition to not being considered in the vaccine given to those who have not completed the schedule or in the boosters for health workers. As shown in Figure 5B, only older adults are protected for measles in all states of the country, with seroprevalence levels in the other age groups below the recommended levels to ensure the elimination of this disease.

**Figure 4.**
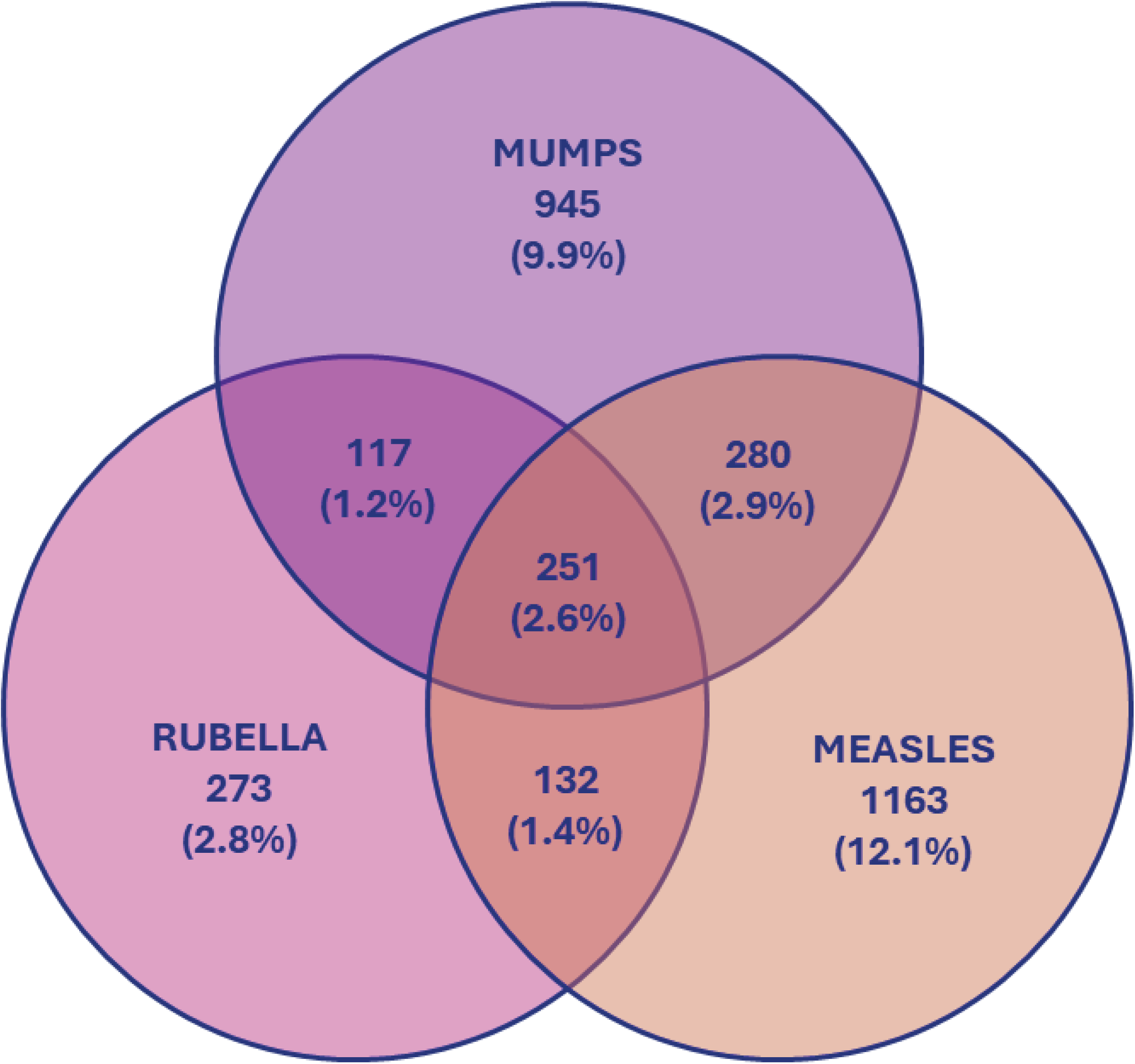
Analysis of seronegativity by combination of diagnoses. Sets of samples negative for IgG against one, two, or three viruses, as well as their intersections. The total numbers of samples negative for mumps, rubella, and measles were 1, 593 (16.6%), 773 (8.1%), and 1,826 (19.0%), respectively. The total numbers of borderline samples for mumps, rubella, and measles were 184 (1.9%), 773 (3.3%), and 256 (2.7%), respectively.

**Figure 5.**
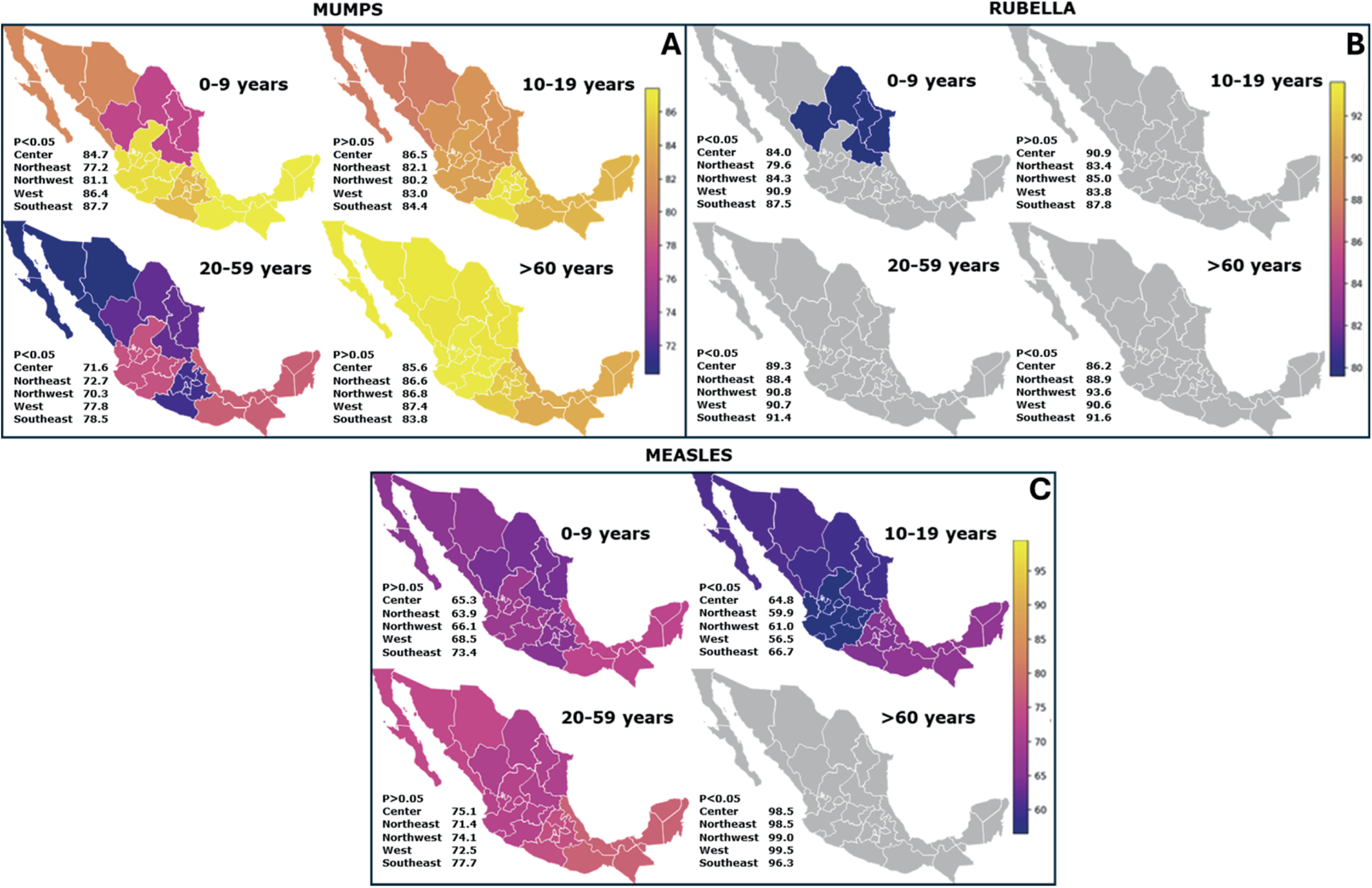
Seroprevalence of mumps, rubella, and measles in Mexico by age group and geographic region. The regional distribution of seroprevalence is shown for: A) mumps, B) rubella, and C) measles. In all panels, regional seroprevalence levels are represented by color scales, with lighter tones indicating greater protection and darker tones indicating lower coverage, whereas regions displayed in gray correspond to areas that achieved the WHO-recommended thresholds required to ensure viral elimination.

To jointly evaluate the three agents contained in the MMR vaccine, samples that tested negative for IgG against one, two, or three viruses, as well as their combinations, were analyzed using intersections. When the cases negative for a single disease were analyzed, the highest percentage corresponded to measles (12.1%), followed by mumps (9.9%) and rubella (2.8%). These percentages were much lower than those observed for overall seronegativity for each disease (21.7%, 18.5%, and 11.4%, respectively).

The intersection between the sets representing the agents included in the double viral vaccine (measles and rubella) was only 1.4%, and it was 2.6% in the case of the agents included in the triple viral vaccine (Figure 4).

## Discussion

In recent years, the seroprevalence of measles, rubella, and mumps has varied significantly worldwide. In developed countries, historically, seroprevalence has tended to be high, as in the case of the United States: 92% for measles, 87.6% for mumps, and 95.3% for rubella (22). However, in other countries, such as Thailand, following the COVID-19 pandemic, seroprevalence was reported to be 77.5% for measles, 55.1% for mumps, and 84.1% for rubella (23). In contrast, a study conducted among healthcare professionals in Spain found seroprevalence of 66.5% for measles, 84.6% for mumps, and 89.8% for rubella (24). All these studies reported significant differences in immunity among young adults. In our study, the overall seroprevalence was 78.3% for measles (95% CI: 77.4%–79.1%), 88.6% for rubella (95% CI: 87.9%–89.2%), and 81.5% for mumps (95% CI: 80.7%–82.2%). Therefore, despite corresponding to the vaccination coverage suggested by the WHO for the elimination of these viruses (MVH: >95%; RVR: 83-86%; OVP: 88-92%) (6-9), our results are only below those of the United States, but with similar findings regarding differences in seroprevalence among age groups.

Countries such as Italy, Greece, Belgium, Croatia, and Hungary usually present high seroprevalence of measles (approximately 89–97%) (25-27), whereas in low- and middle-income countries such as Haiti and the Democratic Republic of the Congo, the seroprevalence of measles is lower (approximately 74–87%) (28,29). Globally, only 65 of the 194 WHO member countries achieved coverage equal to or above 95% in 2022 (30). The most recent study conducted in Mexico to determine the seroprevalence of measles used 3,138 samples (obtained during 2022) and reported two seroprevalence values from the same samples: one obtained by PRNT (82.4%) and the other by ELISA (61.1%) (31). Although our study analyzed measles seroprevalence in Mexico two years later, the values we obtained were much closer to those detected by PRNT, as performed by Carnalla et al. (29).

We opted for CLIA because of its higher sensitivity than that of the ELISA technique (32,33). A study conducted by Brady et al. (34) in 2024 suggested that ELISAs lose even more sensitivity when antibodies induced by vaccination are detected several years postimmunization. Although PRNT assays are still preferred because of their well-known high sensitivity, their large-scale use (as would be required for the more than 9,500 samples analyzed in our study) presents many drawbacks, such as high assay cost, the need for highly trained personnel, and specialized biosafety facilities. Therefore, the high sensitivity of CLIA, combined with the large number of samples analyzed, their distribution across nearly the entire national territory, and the simultaneous identification of IgG antibodies generated against the three agents present in the MMR vaccine, represented the best approach to estimate the real values in the population without the need for viral culture.

Age subgroup analysis for measles and rubella revealed that adult populations exhibited higher antibody titers, especially those who lived during periods of active virus circulation, and therefore may have developed “hybrid” immunity, which is defined as the protection resulting from the combination of a natural infection plus vaccination (35-37). In a cohort study conducted by our research group on the effectiveness of five SARS-CoV-2 vaccines, people with hybrid immunity had higher and longer-lasting antibody titers than those who were only vaccinated (38). Greater protective effects attributed to immunity generated by natural infection than from vaccination have been reported for measles, rubella and mumps (39-41).

The effectiveness of current vaccination regimens for measles might be supported or overestimated because of the hybrid immunity found in adults over 60 years old. However, it is possible to foresee a future in which vaccination alone will be responsible for seroprevalence. This would result in antibody levels 50% lower than those produced by hybrid immunity, which could eventually lead to more frequent outbreaks. This phenomenon was observed in Japan, where even though the seroprevalence increased after measles was eliminated (due to vaccination), there was a decrease in antibody titers in the absence of natural exposure (42).

It has also been reported that, compared to newborns of mothers who had natural infection, newborns of mothers whose immunity comes from vaccination have lower and shorter-lived antibody titers, which could leave the babies vulnerable before they receive their first dose (37). A cohort study carried out in Spain with 146 newborns followed for one year reported that although 91% of newborns had positive antibody titers against rubella, only 13% maintained protective levels at three months, and at nine and twelve months, no protective antibodies were detected (43). Another study conducted in Sri Lanka also demonstrated that immunity to mumps, rubella, and measles transmitted from mothers to newborns decreased considerably during the first 12 months, with seroprevalences of 23.1%, 15.4%, and 9.6% at 6 months for measles, mumps, and rubella, respectively, decreasing to 0% for all three diseases by 12 months of age (44).

This observation highlights the importance of determining the protective antibody titer against measles in order to have more information and analyze the possible need to administer booster doses, especially considering recent reports of outbreaks of measles, rubella and mumps among vaccinated individuals (37).

Mumps differs from measles and rubella not only because its causative agent is the only one among these three diseases that circulates frequently every year but also because, according to the current vaccination schedule, it is not included in the vaccine administered to people who did not complete the two-dose MMR schedule or to healthcare personnel at risk of exposure, as these boosters consist of the MR vaccine (rubella + measles). This suggests that the cases that still occur are partly responsible for slowing the decline in antibody titers observed among the 0–29-year-old group compared with the 40–100-year-old group, reflecting the combined effects of vaccination and accumulated natural exposure.

Seroprevalence analysis by region revealed significant variation among the different regions of Mexico, suggesting heterogeneity in vaccine coverage or exposure to the viruses.

Unexpectedly, adults had the lowest seroprevalence values for mumps, mainly in the northeast, northwest, and central regions of the country. It is concerning that the most susceptible population is in border regions and/or areas with the highest movement of people, especially considering that the most severe cases of mumps usually occur in adults and that there is a high risk of complications such as orchitis, oophoritis, mastitis, pancreatitis, encephalitis, and meningitis (45,46).

The morbidity data reported by the General Directorate of Epidemiology (DGE) in recent years confirm our findings (47), as the majority of mumps cases have occurred among adults, followed by children aged 0–9 years. This immune gap detected in adults, combined with the fact that the greatest number of cases is found in this age group, highlights the need to implement booster immunizations at the end of adolescence or the beginning of adulthood.

Other studies have also highlighted the need for booster doses against mumps. A longitudinal analysis conducted by Qian et al. (48) reported that the mumps vaccine provided protection for approximately one decade, indicating that booster doses in adults may be necessary. Additionally, Pang et al. (49) reported a decrease in antibody levels against mumps following the second dose of the MMR vaccine. Saffar et al. (50) reported that seroconversion after the MMR dose is acceptable; however, to maintain group immunity in the medium and long term, complementary immunization activities seem reasonable.

Owing to the determination of IgG antibodies against all three agents contained in the MMR vaccine, it was possible to analyze cases that tested negative for one, two, and/or all three of the viruses evaluated. We observed that in 2.6% of the total samples, no IgG antibodies were detected against any of the viruses, which could indicate that this percentage represents individuals who have not been vaccinated. However, because mumps continues to circulate year after year, and until just over a decade ago there were also many rubella cases, to this percentage of 2.6% of probably unvaccinated people we should add the percentages from the following groups: measles (neg) + rubella (neg) + mumps (pos) (1.4%) and measles (neg) + mumps (neg) + rubella (pos) (2.9%), amounting to 6.9% of people who have probably not been vaccinated. However, this percentage does not correspond to the seronegativity detected for any of the three diseases when analyzed individually (18.5% for mumps, 11.4% for rubella, and 21.7% for measles), suggesting that seronegativity is not solely due to low vaccine coverage but rather the result of several factors, including people not completing their vaccination schedule on time, a greater-than-expected decline in antibody titers after immunization, the need to update the vaccination schedule in a new context of low viral circulation, or even a low efficacy of the administered vaccine.

Although the results of this study considered samples from across the country, a limitation to consider is that the samples were obtained only from clinical laboratories and blood banks of the Mexican Social Security Institute (IMSS) and therefore represent the population with access to this healthcare system. Extrapolation to the general population should be made with this consideration in mind.

These findings, coupled with the recent increase in measles cases in our country, underscore the need for a comprehensive review of vaccination programs related to these diseases. This review should consider everything from vaccine availability in all regions of the country, the vaccination schedule, and adherence to the schedules in a timely manner, to the effect of the absence of viral circulation on the titer and longevity of the antibodies produced. Furthermore, it highlights the importance of conducting regular serological follow-up studies to keep the country’s epidemiological data up to date.

## Conclusion

The information generated in this study provides a current overview of the country’s immunological status regarding mumps, rubella, and measles, identifying areas for improvement and vulnerable age groups. This report has the potential to guide the design and implementation of strategies aimed at achieving the seroprevalence levels necessary to maintain the elimination of these viruses.

## Acknowledgements

We would like to thank the clinical laboratories (CL) and blood banks (BB) for their participation during the sampling necessary for the completion of this project. In alphabetical order: HGZ No. 1 Aguascalientes (CL and BB), HGZ No. 30 Baja California (CL and BB), HGZ/MF No. 1 Baja California Sur (CL and BB), HGZ/MF No. 1 Campeche (CL and BB), UMAE HE La Raza CDMX (BB), HGZ No. 2 Chiapas (CL and BB), HGR No. 66 Chihuahua (CL and BB), HGZ No. 1 Coahuila (CL and BB), HGZ No. 1 Colima (CL and BB), HGZ/MF No. 1 Durango (CL and BB), UMAE HE 1 El Bajío Guanajuato (CL and BB), HGR No. 1 Guerrero (CL and BB), HGZ/MF No. 1 Hidalgo (BB), UMAE 03 -HESP Oblatos Jalisco (BB), HGO/MF No. 60 México Oriente (CL and BB), HGR No. 220 México Poniente (CL and BB), HGR No. 1 Michoacán (CL and BB), HGZ No. 1 Nayarit (CL and BB), UMAE 34 Cardiología Nuevo León (CL and BB), HGZ No. 1 Oaxaca (CL and BB), HGZ No. 20 Puebla (CL), HGR No. 1 Querétaro (CL and BB), HGR No. 17 Quintana Roo (CL and BB), HGZ/MF No. 1 San Luis Potosí (CL and BB), HGR No. 1 Sinaloa (CL), HGZ No. 2 Sonora (CL and BB), HGZ No. 46 Tabasco (CL and BB), HGR No. 6 Tamaulipas (CL and BB), HGZ No. 1 Tlaxcala (CL and BB), UMAE HE 14 Veracruz Norte (CL and BB), HGZ No. 36 Veracruz Sur (CL and BB), UMAE HESP 1 Yucatán(CL and BB) and HGZ No. 1 Zacatecas (CL and BB).

## Data availability

All data are available in the main text

